# Emergency preparedness planning: lessons from COVID-19 and the opioid crisis in rural Ohio

**DOI:** 10.64898/2026.07.30.26359388

**Authors:** Benjamin R. Westlund, Adams L. Sibley, David C. Colston, Jackson Devadas, Madison N. Enderle, Hannah M. Piscalko, William C. Miller, Vivian F. Go

## Abstract

The COVID-19 pandemic amplified existing public health emergencies, including the overdose crisis.

In rural areas, the effect of emergency preparedness planning on local responses to the pandemic is unknown.

We conducted 52 semi-structured interviews with two participant groups (28 people who use drugs [PWUD] and 24 local stakeholders) in two rural counties in southern Ohio. Interviews explored service disruptions, perceptions of the pandemic, organizations’ pre-existing emergency plans, and communication channels. Transcripts were analyzed thematically to identify data patterns within and between stakeholders and PWUD.

Emergency preparedness plans were often inadequate for the needs of PWUD. Several stakeholders reported that their organizations’ existing plans required updates and additional PWUD perspectives to respond effectively to pandemic-related needs. Both PWUD and stakeholders described widespread service disruptions, including reduced hours, understaffing, and limited access to supplies.

Communication was inconsistent, contributing to challenges in maintaining trust and ensuring access to accurate information. Heightened stigma and law enforcement protocols further discouraged the use of essential services. Despite these challenges, PWUD and stakeholders showed resilience and identified community-driven solutions, including peer distribution networks and rapid transition to telehealth services to mitigate these harms.

The COVID-19 pandemic exposed critical gaps in existing emergency preparedness plans for PWUD in rural Ohio. Ensuring the continuity of essential services, strengthening communication channels, and integrating PWUD perspectives into future emergency preparedness plans are essential to improve responses to future emergencies.

## Introduction

The COVID-19 pandemic intensified the ongoing opioid overdose crisis in the United States by disrupting health systems [1,2], creating social isolation [3], and severely limiting access to health care and social services [4–6]. In 2021, during the pandemic, overdose deaths reached an all-time high of more than 106,000, driven in part by these disruptions and the increasing prevalence of synthetic opioids such as fentanyl [7]. Overdose deaths were especially common in rural areas, rising more rapidly than in urban settings and compounded by limited service infrastructure meant to reduce such deaths [8]. Together, the COVID-19 pandemic created an environment that limited access to essential resources for people who use drugs (PWUD), placed additional strain on already overburdened public health systems, and worsened existing harms.

The pandemic also highlighted the value and limitations of emergency preparedness planning for PWUD. In the United States, preparedness efforts have traditionally focused on natural disasters or terrorism-related events [9], while prolonged public health crises affecting marginalized populations including PWUD have received less attention. Understanding how the pandemic disrupted substance use and harm reduction service systems is essential for improving future preparedness.

The opioid pandemic poses particular challenges for rural communities, including higher opioid prescribing rates, fewer treatment providers, and limited harm reduction resources, such as syringe service programs and naloxone distribution [10–13]. Ohio has been at the epicenter of the opioid crisis, with overdose deaths increasing from 1,539 deaths in 2013 to 4,369 deaths in 2021 [14]. Southern Ohio (generally defined as the counties bordering or proximate to the Ohio River) has experienced a disproportionate burden: four of the five Ohio counties with the highest overdose death rates between 2017 and 2021 are located in that region (Scioto, Gallia, Lawrence, and Pike counties) [15,16]. The overdose rate is exacerbated by limited access to Medications for Opioid Use Disorder (MOUD) and behavioral therapy in much of southern Ohio [17].

In rural Ohio and elsewhere in the United States, the expanding opioid overdose epidemic and the COVID-19 pandemic collided. How local agencies and the PWUD they served managed pandemic-related service barriers remains unclear. Understanding these experiences is essential to inform future preparedness planning.

In this qualitative study, we examined the effects of the COVID-19 pandemic on substance use services and emergency preparedness in two rural counties in southern Ohio. We sought to understand how the pandemic affected drug use, service-seeking experiences, and organizational responses. We also attempted to identify community-driven solutions to inform inclusive and flexible preparedness planning.

## Methods

### Study setting & context

We conducted the study in two rural, Appalachian counties in Ohio that have been heavily affected by the overdose crisis. Like many rural areas, these counties have faced challenges related to healthcare access, overdose mortality, and public health infrastructure; these issues were all exacerbated by the COVID-19 pandemic [14,18]. This research is part of a larger NIH-funded parent study, Implementing a Community-based Response to the Opioid Epidemic in Rural Ohio, one of eight sites in the NIH Rural Opioid Initiative [19,20]. The parent study explored community-level drivers of the opioid epidemic in Appalachian Ohio, with attention to trauma, stigma, and barriers to treatment [21–23]. The present analysis draws on data collected through a COVID-19 supplement to the parent study [24]. This study was approved by the Institutional Review Board of The Ohio State University, serving as the IRB of record, with secondary approval from the Institutional Review Board of the University of North Carolina at Chapel Hill.

### Participants & recruitment

We conducted 52 semi-structured interviews with two participant groups: local stakeholders (n=24) and PWUD (n=28) between June 2021 and February 2022.

Stakeholders included harm reduction workers, public health practitioners, healthcare providers, substance use treatment providers, pharmacists, emergency management personnel, and other community leaders. Stakeholders were eligible if they had at least 2 years of experience providing or supporting health or drug-related services in the study counties. Stakeholders were identified through the Scioto County Health Coalition and the Appalachian Translational Research Network-Ohio (ATRN-Ohio) coalition, and via participant referral.

PWUD participants included men and women who had current or recent injection drug use experience. Participants were selected to capture variation in injection drug use (within the past year), those with longer histories of injection, and some with recent experience accessing health or drug use services. Eligibility criteria included being 18 years or older, residing in one of the two study counties, and reporting injection drug use within the past six months. Participants were recruited using a combination of purposive, convenience, and snowball sampling. Participants were identified through collaboration with local organizations serving PWUD, through peer referrals, and via flyers and business card distribution in community locations. Recruitment was also conducted in community outdoor areas, such as parks and community events.

### Data Collection

Interviews with stakeholders explored: (1) organizations’ pre-existing plans for emergencies; (2) the impact of COVID-19 on the provision of services; (3) strategies to overcome pandemic-related challenges; and (4) barriers to disseminating health information to PWUD.

Interviews with PWUD explored: (1) perceptions of the COVID-19 pandemic; (2) COVID-19 health messaging received and subsequent behavioral changes, such as changes in drug networks and drug transactions; and (3) experiences with stigma, overdose, syringe access, MOUD access, and access to care for infectious diseases (e.g., hepatitis C virus (HCV) and human immunodeficiency virus (HIV)).

Interviews were conducted by two male graduate students in public health, both trained in qualitative interviewing methods, between June 2021 and February 2022, and lasted approximately 45 minutes on average. Interviewers had no pre-existing relationship with participants. Participants were informed that interviewers were conducting research through The Ohio State University and the University of North Carolina at Chapel Hill. All participants were compensated $25 for their time. In accordance with pandemic-related safety protocols, the Institutional Review Board approved an amendment allowing interviews to be conducted virtually via phone or Zoom. All interviews were audio-recorded and transcribed verbatim.

### Data Analysis

All interviews were analyzed thematically in Dedoose using a hybrid inductive-deductive approach consistent with established thematic analysis methods [25]. The codebook was developed using the interview guides for stakeholder and PWUD interviews, as well as through a review of a subset of transcripts to identify other themes. Codes were then organized into a standard qualitative codebook with parent codes representing broader domains and child codes capturing specific subthemes within each domain. The data analysis team met regularly to discuss and revise the codebook before coding and data analysis began.

Three graduate student coders participated in the analysis. To establish intercoder reliability and consistency in coding, the team triple-coded an initial set of three to five transcripts and met to discuss revisions to the codebook. The remaining transcripts were double-coded, and coders continued to meet regularly to discuss discrepancies and ensure reliability. This iterative process identified many patterns across the interviews; however, this paper focuses on the themes most relevant to emergency preparedness planning.

### Ethical Considerations

This study was reviewed and approved by the Ohio State University Institutional Review Board (IRB #2017B0328), which was the primary IRB, with secondary approval from the University of North Carolina at Chapel Hill Institutional Review Board (IRB #20-0660). Written consent initially sought from study participants; however, due to precautions taken during the COVID-19 pandemic, much of the data were collected virtually. In this case, consent was obtained orally, and participants provided verbal informed consent over the phone or video call. The Ohio State University IRB was notified and approved this change in informed consent procedures.

### Artificial Intelligence and Tools

During the preparation of this work, the first author used Claude (Anthropic) to assist with outlining the manuscript and ensuring grammatical accuracy. No new content was generated, and the authors reviewed, revised, and take full responsibility for the final content.

## Results

Data showed that stakeholders and PWUD had to find creative solutions to maintain access to essential services and care due to changes in service delivery caused by the COVID-19 pandemic. Through interviews, five key themes emerged: gaps in emergency preparedness, disruptions to harm reduction and substance use services, breakdowns in communication channels between service providers and PWUD, heightened stigma and law enforcement, and the development of community-driven solutions. Each theme is described in detail below.

## 1. Gaps in emergency preparedness

Stakeholders in both study counties reported that while their organizations did have emergency preparedness plans, they were often outdated and unfamiliar to staff. The plans also did not always consider the needs of PWUD, including plans for disruptions in harm reduction and treatment services. Other stakeholder participants described being unable to locate or describe their organization’s emergency preparedness plans, and several said that the plans were not useful during the COVID-19 pandemic specifically. They noted that protocols were generally oriented towards natural or short-term disasters rather than sustained public health emergencies. Emergency management personnel described feeling ill-equipped for the COVID-19 pandemic. One stakeholder described how the onset of COVID-19 revealed a lack of guidance for coordinating medical or substance use-related issues across sectors:

> *There needs to be a better drawn-out plan for dealing with issues within the community that are out of our resource or asset range. It’s hard to specifically plan for COVID, but we could have a more comprehensive approach that would help enlist different entities within the city. When this hit, it was like—what do we do, who do we call?… Department heads and others in charge of coordination should have a more comprehensive plan for crises like this, especially medical issues, since we’ve already faced the opioid epidemic.” (fire department staff)*

Stakeholders also described feeling confused and frustrated because of overlapping federal, state, and local plans. Stakeholders reported that federal and state directives often superseded existing local plans, limiting their ability to respond effectively to the pandemic, and creating confusion about what counted as “essential services”. As one emergency response coordinator at a local health department said:

> *“We did receive a lot of phone calls asking, you know, who is non-essential and who is essential? So we were trying to help with that definition?…And again, that goes back to that plan that we have, um, and with the state, and so since this was a global pandemic, we didn’t use the local emergency plan, we had to use, we had to follow what the federal government and what the state governments were telling us to do. And so they’re the ones that actually determine what businesses were essential and which ones were not, and gave us the guidelines and then we had to, um, educate our community about how best to proceed.” (health department staff)*

These accounts highlight the challenges that emerged in emergency preparedness planning during COVID-19 and reflect the limitations in local planning and larger structural issues in coordination across federal, state, and local agencies. There were difficulties in turning the federal and state directives into actionable local responses that could respond to the needs of PWUD.

## 2. Disruptions to harm reduction and substance use services

Participants reported that the pandemic disrupted harm reduction services and substance use treatment availability, including facility closures, restricted hours, and limited access to essential supplies. Both stakeholders and PWUD reported that standard pandemic protocols, like social distancing requirements, forced syringe services programs (SSPs) and treatment facilities to limit their operations. These protocols increased barriers to care, particularly for people with limited access to transportation and communication technology.

One participant recalled that the pandemic response forced their local SSP to use appointment-only access:

> *“At first, [the SSP] started doing just one day a week… you had to make an appointment, but they didn’t have a way of advertising it, just word of mouth. The lines were horrible, all the way up to the street. It took like two or three hours to get through.” (person who uses drugs)*

Access to essential harm reduction supplies became difficult during the pandemic, due to the reduced service hours and supply chain issues. As one participant described: *“Nobody had no needles. Everybody did the same ones over and over and over. Everybody’s arms were all messed up. I know me and [redacted name] used the same needles for over a month.” (person who uses drugs)*

In contrast, naloxone access seemingly improved during the pandemic. Some PWUD reported that their local health departments and harm reduction coalitions increased its availability, allowing each person to obtain multiple kits and share them with others. *“Before, you could only get one [Narcan]. Now you can get as many as you want. I take them and go everywhere I go. I usually try and leave a Narcan.” (person who uses drugs)*

There were also changes in service delivery that impacted participants’ access during the pandemic, with some organizations adopting telehealth and drive-through options to preserve access to care. Several PWUD, however, had mixed reactions to these changes, describing them as impersonal or inadequate, especially for group counseling and addiction support.

> *“…we used to meet for groups in person for four or five hours. After the pandemic started, they moved everything to Zoom calls and phone conversations. It just became ineffective—I started using again and pretty much full-on relapsed after that.” (person who uses drugs)*

Others reported a lack of adequate care during telehealth appointments.

> *“When I first went into rehab, I saw a provider in person for my intake. After that, everything was telehealth… I hated it. It’s not that I didn’t feel comfortable, I just didn’t feel like I was getting adequate care.” (person who uses drugs)*

Additionally, fear of contracting COVID-19 discouraged some participants from seeking in-person medical care, even for urgent health problems. *“There’s a couple of times I needed to go, but I didn’t. It was so packed. I just thought, I can heal this up myself.” (person who uses drugs)*

Overall, PWUD reported the pandemic markedly affected local services: (1) reduced services, especially substance use/harm reduction services; (2) decreased availability of harm reduction supplies; (3) changes in care from in-person to telehealth; and (4) avoidance of medical care due to COVID-19-related fears.

## 3. Communication of Information

The pandemic disrupted traditional communication channels between organizations and community members, leading to confusion, mistrust, and reduced access to accurate information about available services and public health guidance. Stakeholders described how in-person outreach and follow-up, previously central to patient engagement, were sharply limited by pandemic-related restrictions. As a result, service providers and public health organizations relied on less effective communication methods such as flyers, social media posts, and mailed letters.

One stakeholder explained the patient follow-up challenges during the pandemic:

> *“Before, we’d do three attempts to get ahold of a positive [hepatitis C] patient. But once COVID got really crazy, all we could do was send them a letter asking them to call us, and most people didn’t [respond]. A lot of that [hepatitis C] education and follow-up just hasn’t gotten done.” (public health nurse)*

The shift away from in-person follow-up also deepened existing inequities with stakeholders reporting that many PWUD lacked stable housing, phone service, and internet access, making digital outreach ineffective. This resulted in organizations relying instead on informal communication channels such as peer networks and word-of-mouth. As one provider noted:

> *“Some of the population we serve…they don’t have a phone, or minutes on their phone, or a permanent address. So reaching those people is hard. Word of mouth tends to work really well in that community.” (public health nurse)*

Some organizations, however, were able to leverage social media as a means of reaching a broader audience, especially among populations with better phone access. A stakeholder from a large health system noted: *“Our biggest avenue during the pandemic was Facebook Live sessions. Our CEO took live questions, which helped us get information out and hear what people were worried about.” (laboratory manager)*

While some were able to access social media, many PWUD continued to face barriers and reported uncertainty about service delivery due to limited phone and internet access. One participant called for *“more awareness put out there for people that don’t understand what’s going on, because there are those that don’t have internet, that don’t have TV, and don’t have the newspaper.”* These gaps disrupted access to care and caused frustration and a feeling of disconnect from the services PWUD relied on.

## 4. Stigma and Law Enforcement Responses

The COVID-19 pandemic created an environment that worsened pre-existing stigma and increased PWUD exposure to the criminal legal system. Social distancing policies designed to reduce viral transmission meant that SSPs limited building occupancy, forcing PWUD to wait outside on the street, where parole officers would sometimes drive by during service hours in an effort to identify and arrest people on probation. One participant explained the risks created by this visibility:

> *“[The syringe exchange] is open on Thursdays. Thursdays is round-up days for probation and like, really? And they make you stand outside because of COVID, so if probation rolls up and there’s somebody on probation, they’re gonna arrest them.” (person who uses drugs)*

Participants described dealing with this by warning others when probation officers were nearby, underscoring how COVID-era public health measures and policies often lead to the creation of new strategies among PWUD to ensure sustained and safe access to harm reduction services.

Despite its life-saving potential, participants also described being fearful of carrying intramuscular naloxone, due to law enforcement perceiving the syringe as being used for illicit drugs.

A community stakeholder noted: *“A lot of people are hesitant to deal with that form of Narcan because you have to pull it up in a needle… and folks would be scared of getting caught with it, especially on probation.” (harm reduction staff)*

This hesitancy was made worse by Ohio’s Good Samaritan Law. As of 2025, this law does not protect people who are on probation or parole in overdose situations [26]. Additionally, the law does not clearly cover intramuscular naloxone possession before an overdose occurs [27], meaning that those most likely to witness or experience an overdose were often least protected by the law.

## 5. Community-Driven Solutions

Despite the systemic challenges and gaps created by the pandemic, PWUD and community stakeholders came up with necessary and novel community-based strategies that helped sustain existing services and support during the pandemic.

Many PWUD took active roles in protecting their communities and became peer distributors by stockpiling extra Narcan and sharing it with other PWUD. One participant explained: *“Yes, they give me as many [Narcan] as I need. I take them and go everywhere I go. I usually try and leave a Narcan. That way I know there’s two shots there.” (person who uses drugs)*

Other PWUD participants ensured that peers had access to harm reduction supplies when SSPs became harder to access. When the syringe exchange closed at the beginning of the pandemic, some PWUD learned through peer networks that certain pharmacies had begun selling syringes and began purchasing and distributing the supplies to others in their networks. *“Now I just go buy a box about once a week. A box of a hundred. And I give them to whoever would want them”* (person who uses drugs)

This example of informal distribution, which emerged as a result of SSP closures, reflects the resilience of PWUD during the pandemic to maintain the safety of themselves and their peers.

Stakeholders and organizations also described rapidly shifting their service models to adapt to pandemic restrictions by quickly introducing telehealth and drive-through models, allowing them to maintain care under otherwise difficult conditions. One provider described: *“We went full telehealth to reduce exposure and risk to our clients and our staff… We would’ve put a lot of clients out on the street, homeless, nowhere to go…”(behavioral health counselor)*

This shift in service delivery allowed clinics to reduce the risk of exposure for their patients and staff.

However, despite telehealth being seen as an adequate solution by clinics and providers, PWUD experiences with this new mode of delivery were often met with dissatisfaction. *“I hated it… It’s not that I didn’t feel comfortable, I didn’t feel like I was getting adequate care.”* (person who uses drugs)

The disconnect between provider intent and client experience reflects a broader tension that arose during the pandemic where solutions designed to maintain access did not always provide adequate care for people in need.

## Discussion

In this qualitative study, we examined the impact of the COVID-19 pandemic on substance use services and emergency preparedness in two rural counties in southern Ohio. Stakeholders indicated that existing emergency plans did not fully address the needs of PWUD, leaving important gaps in services during the pandemic. Both stakeholders and PWUD reported disruptions in service, inconsistent communication, increased stigma, and vulnerability to police surveillance, which may have led to more overdoses and other harms. But PWUD and stakeholder participants also identified community-driven solutions to address these issues and maintain access to supplies and information during a time of widespread disruption. The experiences of both stakeholders and PWUD indicate a need for more flexible emergency preparedness planning explicitly incorporating PWUD’s needs.

In rural southern Ohio, PWUD and providers experienced service disruptions, staffing shortages, and supply limitations but employed creative, community-driven strategies to sustain care and reduce harm. These challenges during the COVID-19 pandemic are similar to those observed elsewhere [6]. The disruptions created a riskier environment for PWUD [2,5,28]. In rural Ohio, PWUD took on informal peer distribution roles, stockpiling and sharing naloxone and syringes with others in their networks. This resiliency mirrors findings from New York City, where syringe service program closures and staffing shortages led PWUD to develop overdose-prevention strategies, such as forming living “bubbles”, taking turns using, and stockpiling supplies when available [29]. But the peer distribution described by PWUD in Ohio did not occur through syringe exchanges, which operated under Ohio’s commonly used 1:1 syringe exchange model [30]. Instead, the effort emerged as a workaround when health departments arranged for local pharmacies to sell syringes. While this approach prevented further harm to PWUD, it also highlights the shortcomings of a 1:1 exchange model in meeting PWUD’s needs during emergencies. Needs-based syringe service programs, where participants can receive as many syringes as they need, have been shown to reduce syringe sharing and decrease HIV incidence in Appalachian settings [31,32]. Transitioning to a needs-based model could reduce reliance on peer-distribution networks during future emergencies and better serve PWUD under normal operating conditions.

Building on this resilience among PWUD, providers and stakeholders transitioned to telehealth and implemented drive-through care models to maintain continuity of care. In Chicago, transitioning to telehealth, implementing hybrid-care models, and developing methods to deliver harm reduction supplies through mobile or community-based approaches was a critical response [33]. Yet, we found that the shift to telehealth was sometimes inadequate for PWUD in rural Ohio. As seen in prior work [34], access to telehealth was itself a barrier for some, given limited phone and internet connectivity in rural areas.

Among those who did access it, PWUD described telehealth as impersonal and insufficient, with some feeling they were not receiving the level of care they needed. This disconnect between clients’ experiences and well-intentioned providers and stakeholders points to the limitations of telehealth as a complete replacement for in-person services.

PWUD discussed their reliance on word-of-mouth for receiving pandemic-related information, struggling with phone and internet access, and missing medical care due to a lack of follow-up from organizations and a fear of contracting COVID. In other rural contexts, PWUD also often lacked reliable internet access, relied on informal methods of receiving COVID-19 information, and often avoided seeking medical care due to stigma and mistrust of medical providers [1]. We found these challenges were exacerbated in a rural context where peer-driven communication networks were often the primary means of disseminating information during the pandemic.

Stigma and law enforcement responses during the COVID-19 pandemic exacerbated existing inequities among PWUD in southern Ohio. In urban areas, such as Toronto, heightened stigma and policing during the pandemic discouraged PWUD from accessing harm reduction services [35]. Similarly, participants in southern Ohio described that having to wait outside of SSPs on the street increased their exposure to police and parole officers, deterring some from accessing harm reduction services altogether. Fear of arrest, stigma from first responders, and negative perceptions of the local judicial system were primary barriers to calling 9-1-1 during an overdose in the same rural Ohio counties [36]. The limited uptake of overdose prevention tools like naloxone among law enforcement further underscores how gaps in coordination between public health and criminal legal systems exacerbated risks for PWUD during a public health emergency [35,36].

We found systemic emergency preparedness planning shortcomings, such as conflicting federal, state, and local directives and the exclusion of harm reduction experts and PWUD from preparedness efforts.

Systemic planning shortcomings exacerbated pandemic-related barriers. Without more inclusive and coordinated preparedness planning, future public health emergencies will be insufficiently addressed and may deepen existing inequities. Strengthening preparedness requires integrating the perspectives of harm reduction experts and PWUD. Emergency preparedness plans must also address harm reduction and treatment services.

Drawing on perspectives of community stakeholders and PWUD, we identified several opportunities to strengthen future preparedness planning (Table 1). The recommendations highlight broader implications for public health practice, emphasizing the need for coordinated, inclusive, and adaptable planning that prioritizes harm reduction, communication, and continuity of services during public health emergencies.

**Table 1.** Recommendations for Emergency Preparedness Based on Findings.

| **Domain** | **Key Findings** | **Recommendations** |
| --- | --- | --- |
| *Planning & Coordination* | Emergency plans did not address PWUD; conflicting directives created confusion | Ensure that harm reduction and treatment providers are incorporated into county and state emergency preparedness planning, with flexible protocols that can be adapted to local contexts [5,9,28,37]. |
| *Communication of Information* | Digital divide and mistrust limited information access; reliance on word-of-mouth | Use multipronged strategies: peer-based outreach, flyers, radio, and social media to reach diverse audiences [1,6,29,33]. |
| *Substance Use and Harm Reduction Services* | SSPs and treatment centers reduced or closed; telehealth was often inadequate | Codify harm reduction and treatment services as essential during emergencies and ensure continuity through flexible, low-barrier service delivery models [2,12,34,38,39]. |
| *Law Enforcement and Legal Protections* | Outdoor SSP waiting lines increased visibility to police; fear of arrest and naloxone- | Align law enforcement practices with harm reduction during emergencies by reducing |
|  | related stigma deterred service use | punitive enforcement and ensuring officers are equipped to respond to overdoses. SSPs and harm reduction programs can also adopt low-barrier operational measures (e.g., private entrances, scheduling adjustments) to reduce PWUD's visibility and exposure to surveillance while accessing services [13,35,36,40]. |
| *Supply Management* | Severe shortages of essential supplies. PWUD stockpiled/distributed supplies; peer networks filled service gaps | Strengthen emergency supply chains for harm reduction materials and integrate peer networks into distribution planning [29,31,32,34]. |

Our findings underscore the need for more coordinated and responsive emergency preparedness planning that addresses the needs of PWUD. Participants described extensive service interruptions and communication challenges during the COVID-19 pandemic, while also identifying practical solutions to inform future crisis planning. The recommendations span five domains: emergency planning, communication, substance use and harm reduction services, law enforcement, and supply management.

Local emergency planning operations did not adequately account for infectious disease emergencies beyond bioterrorism [9] and rarely incorporated input from substance use treatment and harm reduction services. Effective planning must reflect the realities of substance use and include the perspectives of PWUD and service providers. Other places have developed infectious disease emergency response plans that explicitly define cross-agency roles, outline procedures for maintaining essential services, and integrate harm reduction into preparedness frameworks [37]. Adapting elements of such approaches could strengthen local coordination and ensure that harm reduction and treatment services are incorporated into county and state preparedness efforts.

Disruptions to traditional communication channels left many PWUD uncertain about service availability and access. Preparedness plans must therefore include multilevel communication strategies that reach individuals without reliable phone or internet access. Combining digital platforms with low-tech approaches, such as printed materials distributed through harm reduction kits and peer-based outreach, is critical. While existing resources may offer general guidance, they must be paired with strategies that reach the most disconnected community members to ensure equitable access to information during sustained emergencies.

Service disruptions and closures during the pandemic limited access to sterile supplies, MOUD, and other essential services. Future preparedness efforts should explicitly designate harm reduction and substance use treatment as essential services at the state and local levels. Continuity of care may require flexible service models such as mobile or drive-through distribution and telehealth, while accounting for limited phone and internet access among PWUD. Integrating peer networks into service and supply distribution can also strengthen outreach and access.

Participants described how pandemic-related restrictions increased their exposure to police and probation officers, discouraging use of harm reduction services. Emergency preparedness planning should better align law enforcement practices with harm reduction principles during public health crises. This means prioritizing public health over punitive enforcement of low-level offenses and ensuring that officers are equipped and trained to respond to overdoses with naloxone. Limiting unnecessary criminal-legal contact during emergencies may reduce harm and improve coordination between public health and public safety systems.

Participants consistently reported shortages of essential supplies, including syringes, naloxone, and basic protective equipment. It’s important for emergency preparedness planning to focus on strengthening supply chains at the local level and preserving access to harm reduction materials during emergencies. While backup distribution pathways should be established, peer distribution networks, which already play an important role, should also be integrated into emergency planning. It is essential to build resilient, flexible, and community-driven supply systems to maintain access to lifesaving resources during prolonged and overlapping public health emergencies.

Together, these recommendations offer practical guidance for future emergency preparedness efforts. Incorporating the perspectives of PWUD and harm reduction providers, using both digital and low-tech communication strategies, and aligning law enforcement and public health priorities may help communities better protect vulnerable populations during future emergencies.

This study has some limitations. Interviews were conducted in two rural counties in southern Ohio, so findings may not generalize to urban settings or other rural settings with different demographic, infrastructural, or policy contexts. Interviews were conducted between June 2021 and February 2022, so participants interviewed later in this window may have relied on retrospective recall of their experiences early in the pandemic, which could affect the specificity and accuracy of their accounts. Despite these limitations, the inclusion of both stakeholder and PWUD perspectives in a rural setting provides a more comprehensive view and represents a perspective often underrepresented in prior, largely urban and provider-focused research.

## Conclusion

This study showed that the COVID-19 pandemic exposed critical gaps in emergency preparedness planning for PWUD in rural southern Ohio. Disruptions to harm reduction and treatment services, unclear communication, and heightened concerns about stigma and law enforcement responses increased barriers to care at a time when services were most needed. Stakeholder and PWUD participants identified practical strategies such as peer communication networks and collaboration, continuity of essential services, and policy changes to mitigate harms caused by this public health emergency. Improving responses to future crises will require emergency preparedness planning that meaningfully includes

PWUD and harm reduction partners to ensure public health is not compromised. Future research should identify which community-driven harm reduction practices are most effective in helping PWUD during disruptions and evaluate how these strategies function across diverse rural and urban settings, so that planning efforts safeguard PWUD in both routine and crisis conditions.

## Data Availability

The data underlying this study cannot be made publicly available due to ethical and legal restrictions. Participants were recruited from a small, geographically concentrated rural population and disclosed sensitive, potentially legally incriminating information related to illicit drug use during semi-structured interviews. Given the small sample size and specific geographic setting, there is a meaningful risk that de-identified transcripts could still permit participant re-identification within these communities. Public access to de-identified transcripts would violate protections issued by Ohio State University's Institutional Review Board. Approval to access data can be sought from Ohio State's Huron IRB System via email.

## Acknowledgements

We thank the Scioto County Health Coalition and the Appalachian Translational Research Network-Ohio (ATRN-Ohio) coalition for their support with participant recruitment. We are grateful to the local community stakeholders and people who use drugs who shared their valuable experiences and perspectives for this study. We’d also like to thank Abby Spears for her contribution to this work.

